# National Prevalence and Determinants of Concurrent Wasting and Stunting among Under-Five Children in Yemen: Evidence from the 2022–2023 Multiple Indicator Cluster Survey

**DOI:** 10.64898/2026.10.06.26364923

**Authors:** Abdullah Salman, Rifa Sunzida, Bushra Kabir, Rajat Das Gupta

**Author notes:** Corresponding Author : Abdullah Salman, Email address of the Corresponding Author.

## Abstract

**Background:** Concurrent wasting and stunting (WaSt) is a severe form of child undernutrition, linked to higher mortality and long-term negative health outcomes. Malnutrition is a significant challenge in Yemen, where the country is experiencing a long-standing conflict, food insecurity, and socioeconomic deprivation. But there is still limited evidence on the burden and determinants of WaSt in Yemen on a national level. This study aimed to estimate the prevalence and identify factors associated with WaSt among children under 5 years in Yemen.

**Methods:** We conducted a cross-sectional analysis using nationally representative data from the 2022–2023 Yemen Multiple Indicator Cluster Survey (MICS). A significantly large dataset of 17,734 children aged 0–59 months from all the 22 governorates of Yemen was analyzed. Concurrent wasting and stunting (WaSt) were defined as the coexistence of a weight-for-height z-score (WHZ) below −2 standard deviations and a height-for-age z-score (HAZ) below −2 standard deviations from the WHO Child Growth Standards in the same child. Survey-weighted logistic regression was performed to analyze the factors associated with WaSt among the under 5 children in Yemen.

**Results:** The national prevalence of WaSt was 8.4% (95% CI: 7.6–9.2). The highest prevalence of WaSt was found in children aged 12-23 months (12.0%) and in the Western Coastal region (11.6%). After adjustment, male sex (AOR 1.52, 95% CI: 1.30–1.80), residence in the Western Coastal (AOR 2.00, 95% CI: 1.60–2.40), Eastern (AOR 2.00, 95% CI: 1.50–2.70), and Southern regions (AOR 1.60, 95% CI: 1.20–2.10), poorest household wealth (AOR 1.96, 95% CI: 1.30–2.90), moderate (AOR 1.51, 95% CI: 1.13–1.94) and severe food insecurity (AOR 1.94, 95% CI: 1.40–2.60), and recent diarrhoea (AOR 1.22, 95% CI: 1.02–1.44) were associated with higher odds of WaSt. Having a handwashing facility and improved sanitation were protective.

**Conclusion:** At least 1 in every 12 under-five children in Yemen was affected by WaSt, and there were significant socioeconomic, geographic, food-security and WASH-related factors influencing this devastating outcome. Instead of simple, one-dimensional approaches, multi-sectoral interventions are needed to address this adversity. And those efforts must be integrated into the system, interconnected with each other and specifically targeted to vulnerable children and households.

## Introduction

Malnutrition severely disrupts a child’s healthy growth, development, learning, and overall well-being ^1^. Among the under 5 children, undernutrition commonly persists as stunting, wasting and underweight ^2^. Wasting is diagnosed when a child’s weight-for-height z-score falls more than two standard deviations below the WHO growth-standard median, reflecting rapid weight loss or failure to gain weight ^3^. Wasting is therefore known as acute malnutrition ^1^. Stunting denotes low height-for-age — a height-for-age z-score more than two standard deviations below the WHO growth-standard median ^4^ . While wasting is an acute manifestation of nutritional deficiency, stunting represents chronic malnutrition with largely irreversible effects ^5^. Concurrent wasting and stunting (WaSt) is defined as the presence of both wasting and stunting in the same child at the same time point in a population ^6^ . WaSt therefore represents both acute and chronic forms of malnutrition together ^7^. It is now recognized as a distinct, serious form of child malnutrition. Compared with children who are only wasted or only stunted, those with WaSt have nearly 12 times greater likelihood of death ^8^. Existing evidence also shows that stunting and wasting are closely related and more often coexist together in children ^9^. Thus WaSt not only gives rise to various short term complications leading to child mortality, but also causes numerous long-term complications as well. These long term effects include sustained linear growth faltering , delayed neurodevelopment , impaired cognitive functions, frequent ARIs and diarrhea , metabolic and cardiovascular diseases and lack of immunity ^6, 10–11^. Globally, at least 150 million under 5 children are stunted and 43–47 million are wasted ^12^. Based on data from 84 countries it is said that approximately **3%** of children around the world are concurrently wasted and stunted (WaSt), with Africa (3.5%) and Asia (3.4%) leading in the burden ^13^. However, selectively in the LMIC countries, this burden is much higher, especially countries that have ongoing conflict and humanitarian crisis with large number of marginalized populations^6^ . Therefore, estimating the burden of WaSt in these LMICs and their key determinants is highly critical. Only this can unlock ways to identify the communities vulnerable to WaSt in those countries and facilitate the implementation of targeted public health interventions.

Yemen is a conflict-affected low- and middle-income country, where years of war, economic collapse, and a decimated health system have driven extremely high levels of child malnutrition and deprivation^14^. Children in Yemen are disproportionately affected, with around half of all under-fives estimated to be malnourished and hundreds of thousands suffering severe acute malnutrition^15^. A previous study conducted among a marginalized community in Sana’a City, Yemen, reported a WaSt prevalence of 10.7%^16^. However, its small sample size and focus on a single community reduce the external validity of the findings to the broader child population of Yemen. Despite the high burden of child undernutrition in Yemen, there has been no study on nationally representative evidence on the prevalence and determinants of concurrent wasting and stunting. Such evidence is essential to identify high-risk populations and regions and to guide targeted, evidence-based nutrition interventions and policies. In 2022-23, UNICEF conducted a Multiple indicator cluster survey (MICS) in Yemen to generate nationally representative data on the health and well-being of children, women, and households. This study therefore aims to utilize this data from MICS 2022 and evaluate the national prevalence of concurrent wasting and stunting and its associated factors across Yemen. It will provide the first nationally representative evidence on the burden and determinants of concurrent wasting and stunting in Yemen. The findings of this study will help policymakers and public health practitioners identify vulnerable populations, prioritize resource allocation, and design targeted interventions to reduce the burden of concurrent wasting and stunting in Yemen.

## Methods

### Study design and population

The Multiple Indicator Cluster Survey (MICS) in 2022-23 from Yemen provided secondary data for this study ^17^. MICS is a UNICEF-coordinated program of nationally representative household surveys designed to produce standardized, internationally comparable data on the health and welfare of women and children ^18^. To make the survey nationally representative, a two-stage stratified sampling design was employed. Comprehensive details on the survey methodology, sample techniques, and datasets may be found in the Yemen MICS report and the MICS program website ^17^. From the Yemen MICS 2022–23 dataset, records representing 20,200 children were screened for eligibility. After excluding records with no matching maternal data, missing HAZ or WHZ scores, and anthropometric values violating the WHO data quality criteria ^17^, 17,492 children comprised the final unweighted analytical sample. Applying the sampling weight along with stratification and clustering, the weighted analysis represented a survey population of 17,734 children. The complete process of participant selection is shown in Figure 1.

**Fig 1:**
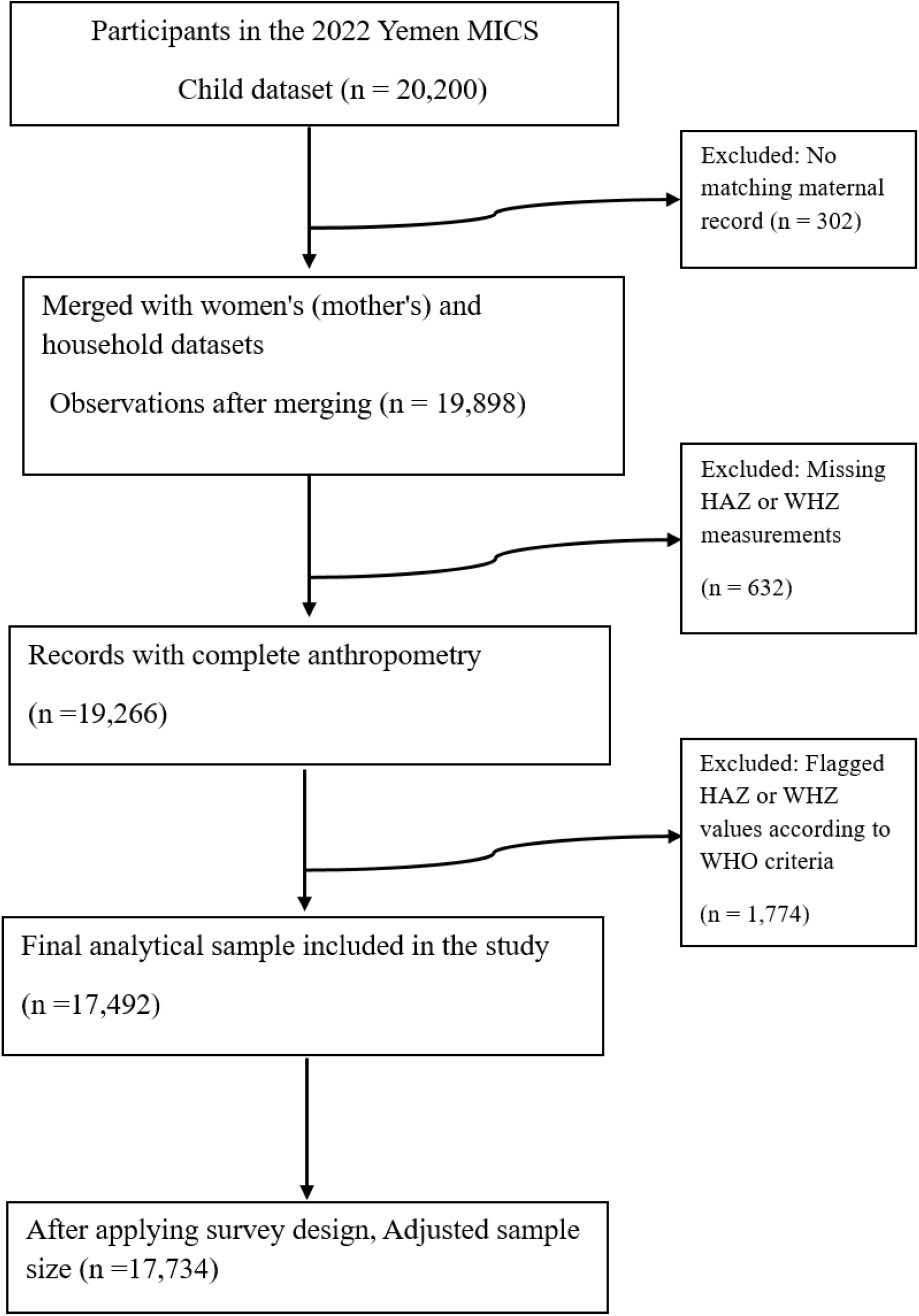
STROBE participant flow diagram

### Study variables

Concurrent wasting and stunting (WaSt) status was the dependent variable in this study, and it was classified into two groups: "Both wasted and stunted" and "Not concurrent wasting and stunting." HAZ score < -2 SD and WHZ score < -2SD were used to define this ^6^. Sociodemographic, household, environmental, and childhood morbidity-related exposure variables were grouped together. Six age groups were created: 0–5 months, 6–11 months, 12–23 months, 24–35 months, 36–47 months, and 48–59 months. Yemen is divided into 22 administrative governorates. Based on their geographic locations, these governorates were split into five regions: Northern, Southern, Eastern, Western Coastal, and Central. Households were considered food insecure when limited money or other resources prevented them from reliably obtaining sufficient, safe, and nutritious food for normal growth and an active, healthy life ^19^. Food insecurity was measured with FAO’s eight-item Food Insecurity Experience Scale, which records increasingly severe experiences during the preceding 30 days — eating less than desired, worrying about food, and going a whole day without eating ^20^. The eight binary items (yes = 1, no = 0) were summed into a 0–8 score, with higher totals indicating greater severity. Households were then grouped as food secure (0), mildly (1–3), moderately (4–6), or severely (7–8) food insecure ^21^.The household wealth index, the mother’s educational level and functional difficulty, whether the family lived in an urban or rural area, and owned domestic or agricultural animals were additional sociodemographic factors. Malnutrition in children is closely linked to recurrent illnesses and infections. Thus, morbidity in children was also evaluated. A recent (within the last two weeks) history of fever and diarrhoea were the childhood morbidity variables. The state of the water source, sanitary toilet, and handwashing area were also considered because diarrhoea is linked to WASH practices. Drinking-water sources and sanitation facilities were classified according to the MICS report’s categories ^18^. Piped water, protected wells and springs, rainwater collection, tanker- or cart-delivered water, and bottled or sachet water counted as improved sources; unprotected wells and springs and surface water counted as unimproved ^22^. Sanitation was graded on the same scheme: flush or pour-flush toilets, pit latrines with a slab, ventilated improved pit latrines, and composting toilets were considered improved, whereas bucket or hanging latrines and pit latrines without a slab were not ^22^.

### Statistical analysis

Descriptive statistics were used to summarize the study population’s characteristics for each of the explanatory variables. The Chi-square test was used to evaluate bivariate relationships between concurrent wasting and stunting (WaSt) and the independent variables. Because the survey used stratified, multistage cluster sampling with unequal selection probabilities, weighted logistic regression was fitted so that point estimates and standard errors reflected the complex design ^23^. There were two stages to the analysis. Initially, the crude relationship between each explanatory variable and WaSt was investigated using univariable survey logistic regression (Model I). Then, by simultaneously entering variables, a multivariable survey logistic regression model (Model II) was developed to estimate adjusted associations. Statistical significance was defined as a p <0.05. The software StataMP version 17.0 was used for all statistical analyses. To demonstrate the study’s geographical setting in relation to the burden of concurrent wasting and stunting, a choropleth map (shown in Figure 2) was made using QGIS software version 4.0.3 .

**Fig 2:**
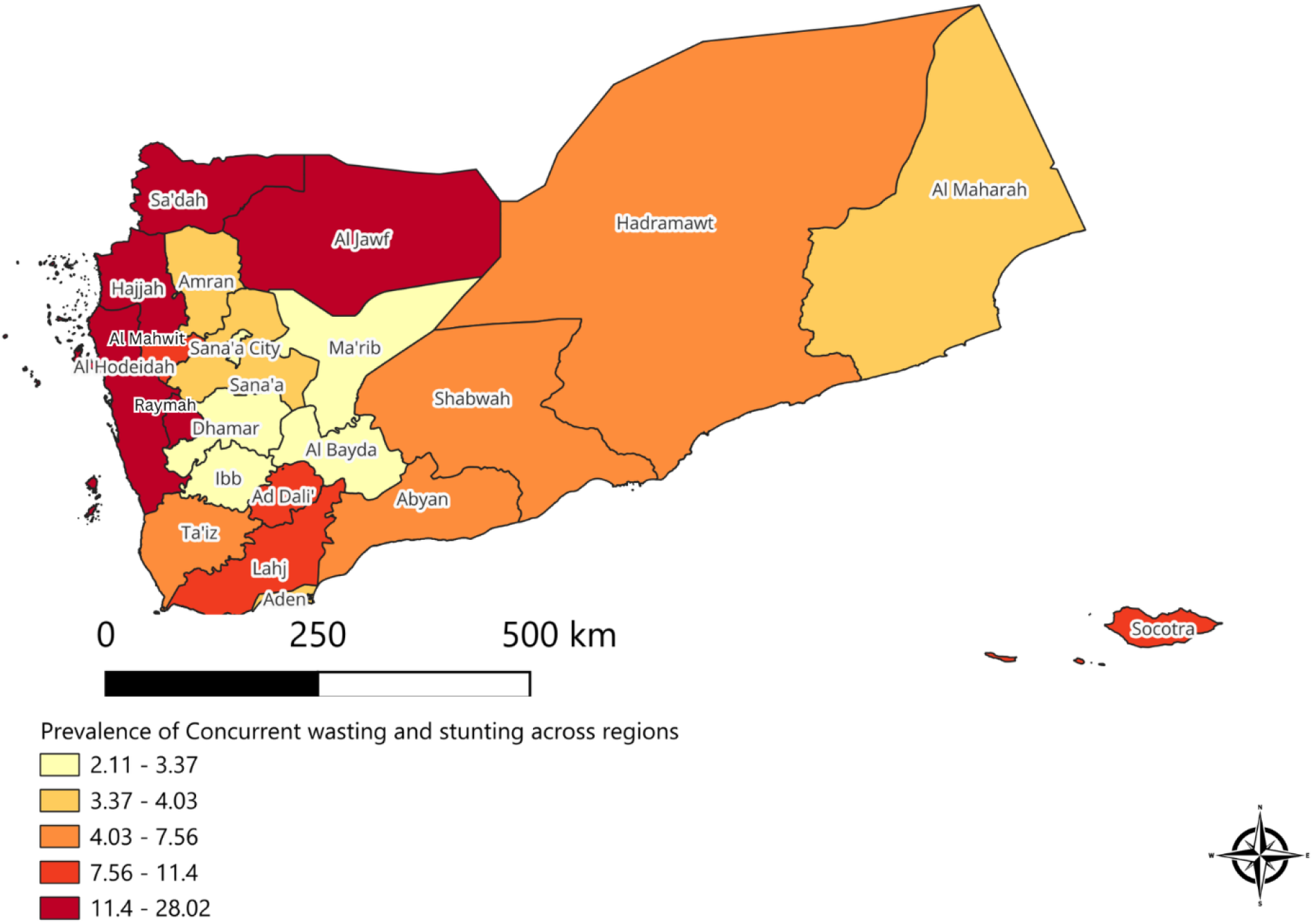
Choropleth map showing the distribution of concurrent wasting and stunting among under 5 children across all the governorates in Yemen

## Results

After excluding records with missing anthropometric data, 17,734 children under 5 years of age were included in the final analysis. The overall national prevalence of concurrent wasting and stunting was 8.4% (95% CI 7.6–9.2), indicating that approximately one in every 12 children under five in Yemen simultaneously experienced acute and chronic undernutrition.

### Distribution of WaSt across sociodemographic, household, environmental, and child health characteristics

The distribution of WaSt across sociodemographic, household, environmental, and child morbidity-related characteristics is presented in Table 1. The sample comprised slightly more male (51.2%) than female (48.8%) children. WaSt prevalence was markedly higher among boys (9.7%) than among girls (7.0%), a difference that was statistically significant in bivariate analysis (p < 0.001). Child age was strongly and significantly associated with WaSt (p < 0.001). Prevalence was highest among children aged 12–23 months (12.01%), followed by those aged 36–47 months (9.04%) and 24–35 months (8.5%), whereas infants aged 0–5 months had the lowest prevalence (3.33%). This pattern suggests that children become more susceptible once complementary feeding is initiated during the early weaning period. WaSt prevalence was significantly higher in rural (9.7%) than in urban (5.0%) areas (p < 0.001).

**Table 1.**
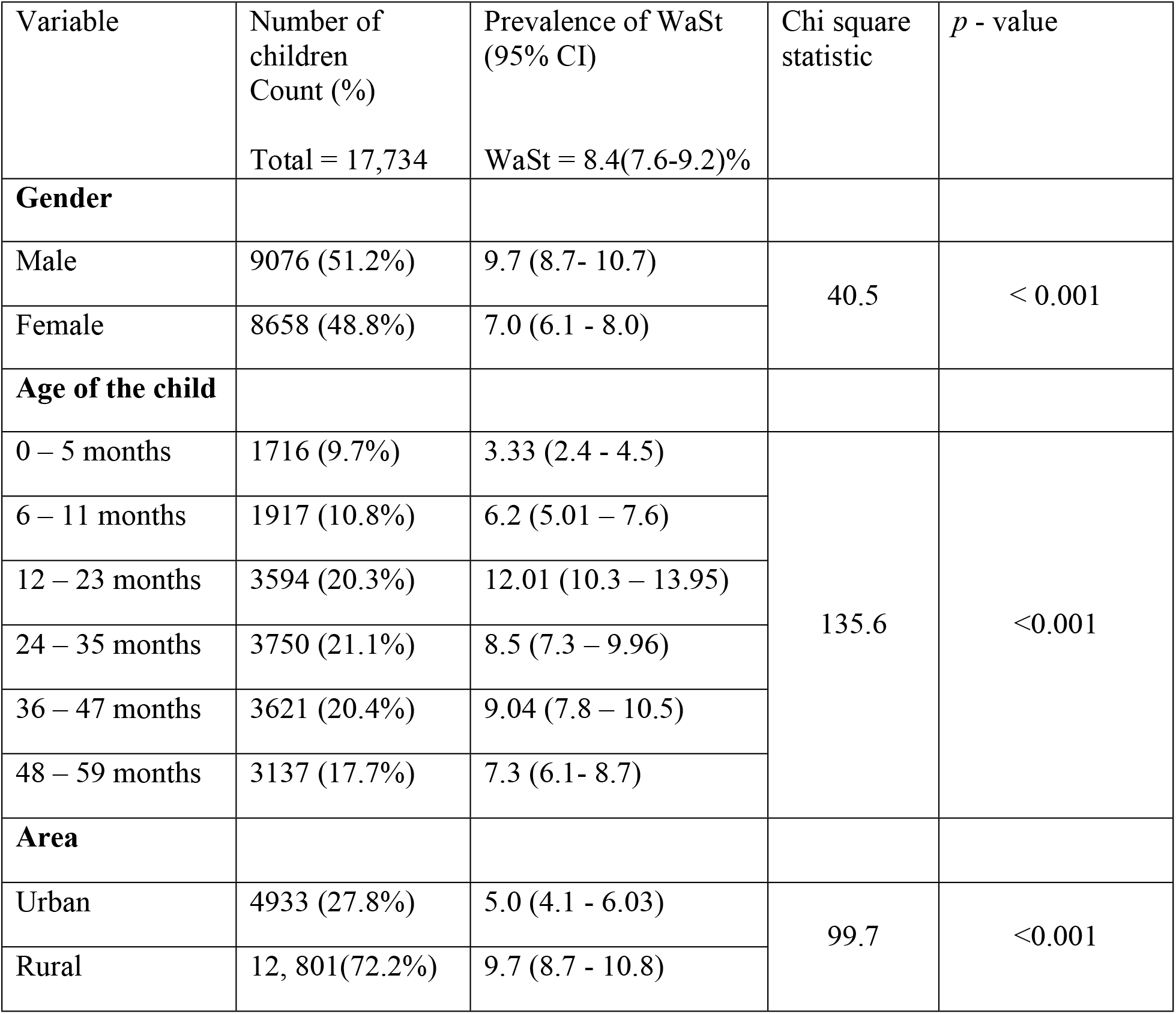
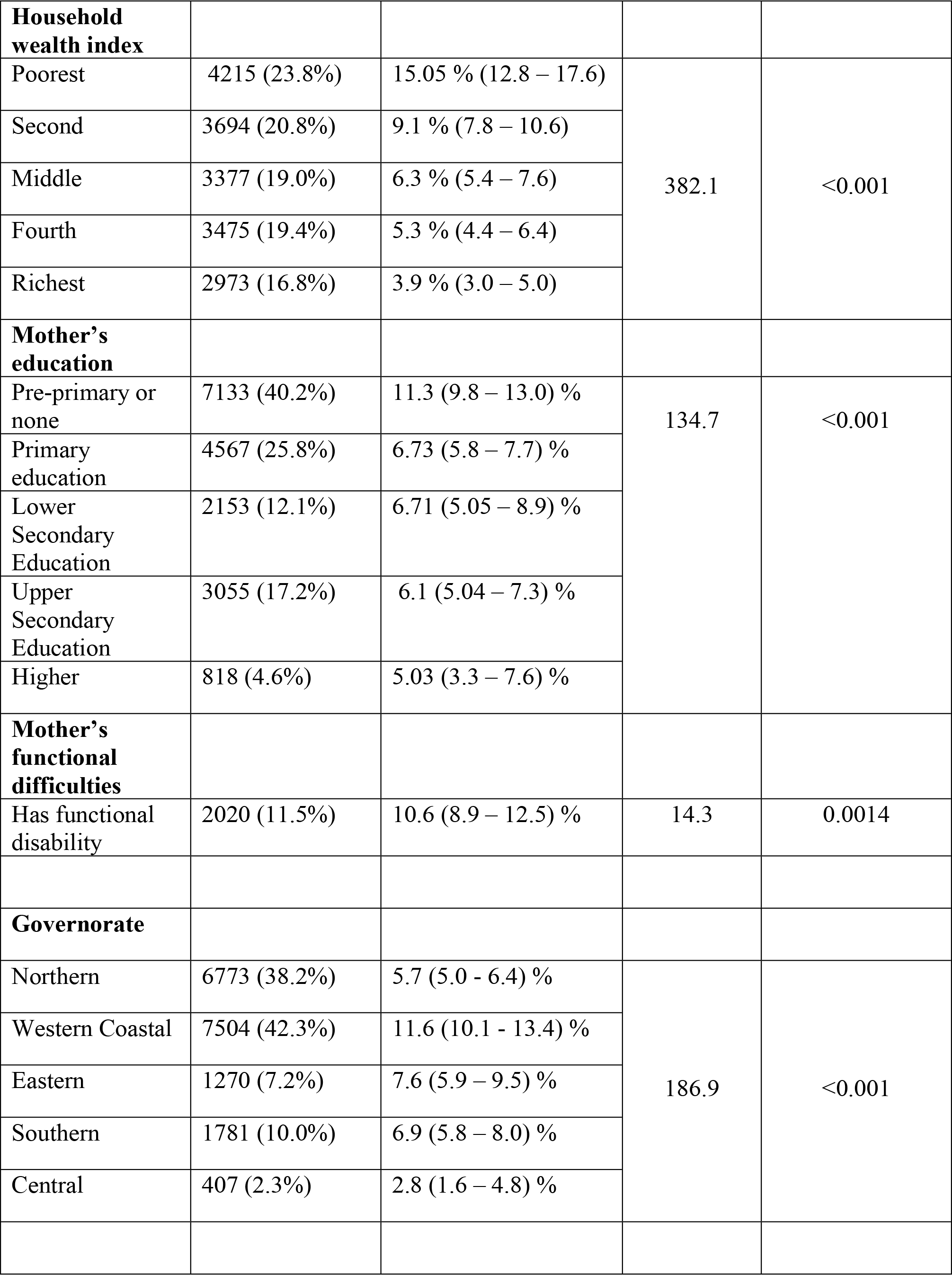
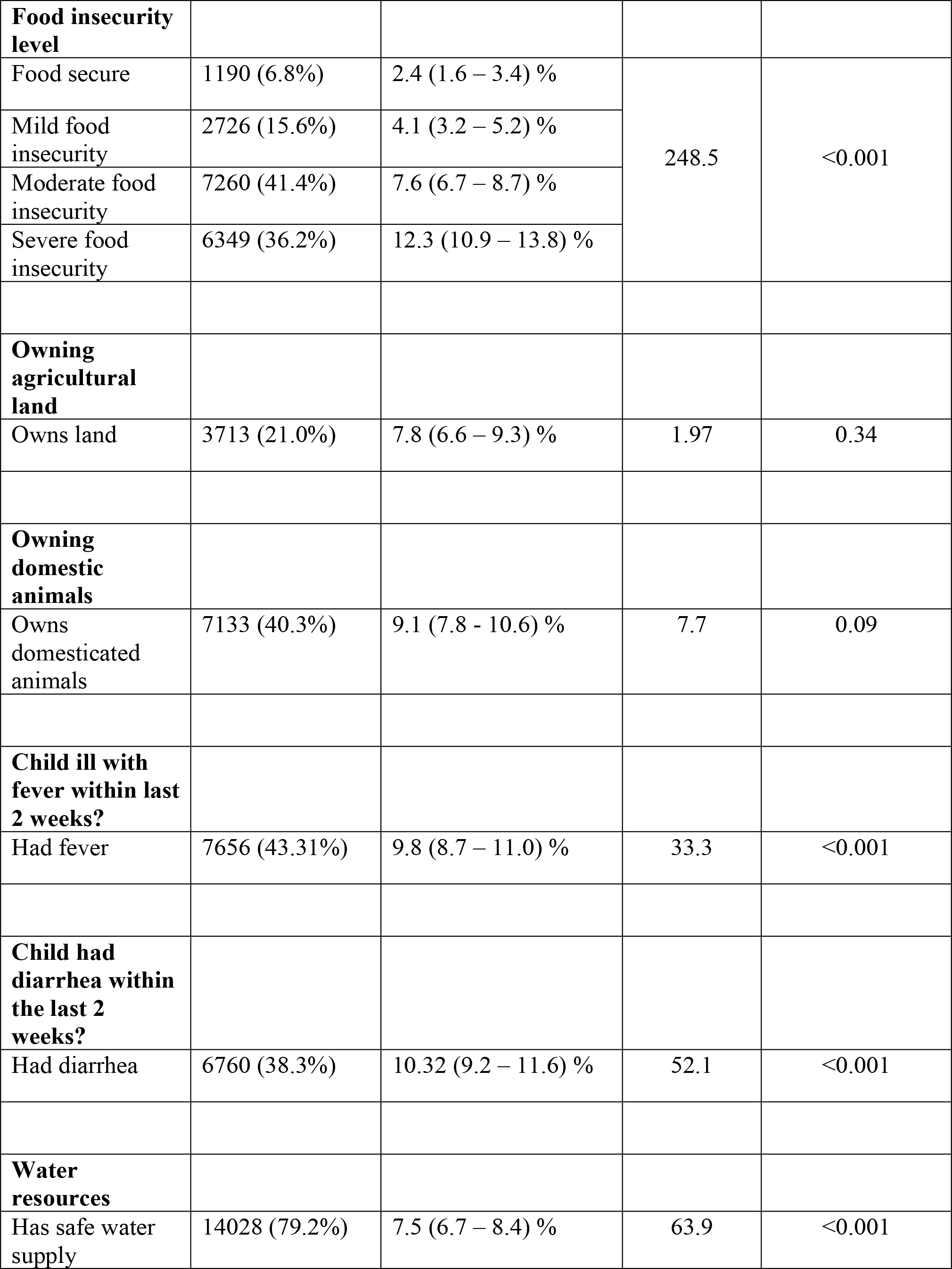
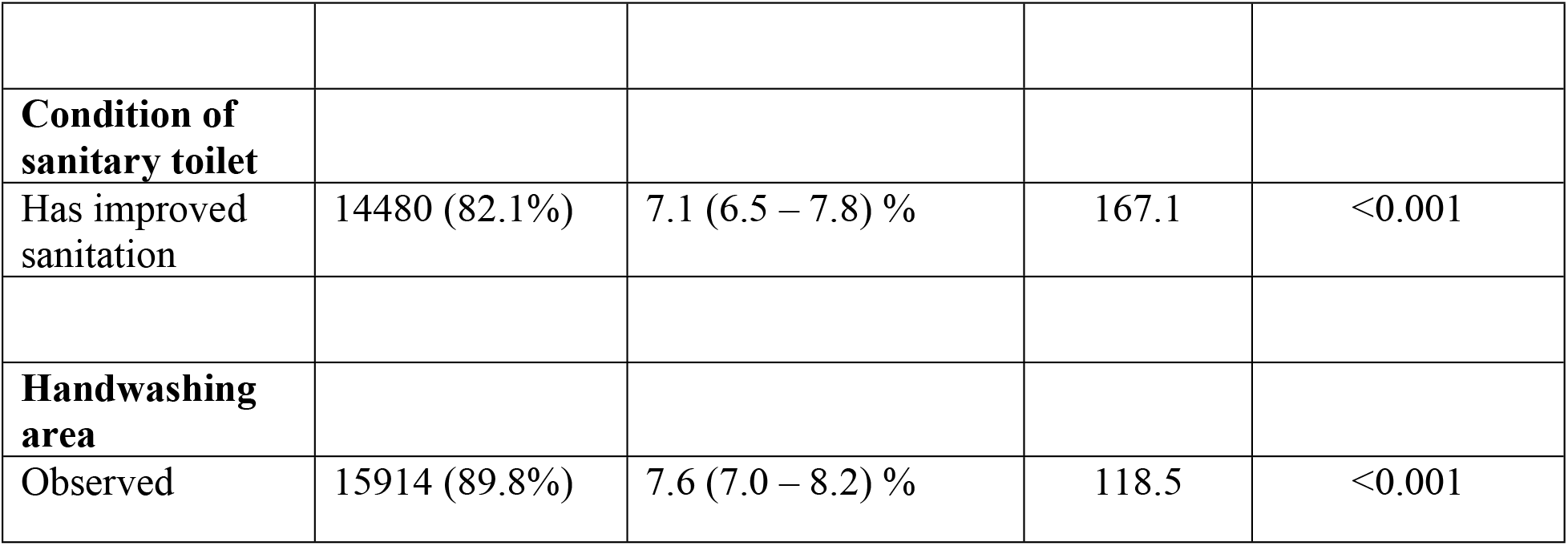
: Distribution of concurrent wasting and stunting across various sociodemographic, household, environmental and child health characteristics among under-five children

Yemen is administratively organized into 22 governorates. Significant geographic heterogeneity in WaSt prevalence was observed at the governorate level (p < 0.001), indicating a high spatial concentration of co-occurring stunting and wasting. WaSt prevalence was inversely related to the household wealth index, and this association was highly statistically significant (p < 0.001): children in the lowest wealth quintile had the highest prevalence (15.05%), nearly four times higher than that of children in the wealthiest households (3.9%). Maternal education was also significantly correlated with WaSt (p < 0.001). Children of mothers with no or pre-primary education had the highest prevalence (11.3%), whereas children of more educated mothers had the lowest (5.03%). The analysis further found that nearly 11.5% of mothers had some sort of disability, and maternal functional challenges were significantly correlated with WaSt among Yemeni children under five (p < 0.01).

Household food insecurity emerged as one of the most prevalent problems identified in the analysis. Nearly 36.2% of children lived in severely food-insecure households, whereas only 6.8% lived in food-secure households. WaSt was significantly associated with household food insecurity (p < 0.001) and became more common as food insecurity worsened: prevalence was 2.4% in food-secure households, 4.1% in mildly food-insecure households, 7.6% in moderately food-insecure households, and 12.3% in severely food-insecure households. Accordingly, WaSt was roughly five times more common in severely food-insecure households than in food-secure households. Domestic animals were present in 40.3% of households and nearly 21% had agricultural land; however, neither factor was associated with WaSt prevalence.

WaSt was significantly correlated with the experience of recent illness, including fever and diarrhoea (p < 0.001). Among children who had a fever within the previous two weeks, 9.8% were both stunted and wasted, while among those who experienced diarrhoea within the previous two weeks the prevalence of WaSt was even higher (10.32%). Because childhood diarrhoea is closely associated with community WASH practices, these factors were also examined. WaSt prevalence was significantly associated with access to safe drinking water, sanitary toilet facilities, and designated handwashing areas (p < 0.001): children from households with safe drinking water sources had a WaSt prevalence of 7.5%, those with improved sanitation facilities a prevalence of 7.1%, and those with designated handwashing facilities a prevalence of 7.6%.

### Spatial distribution of WaSt across Yemen

Alongside sociodemographic and health-related factors, geographic location was another focus of this national-level analysis. WaSt prevalence varied significantly across Yemen’s five geographical regions (p < 0.001): it was highest in the Western Coastal region (11.6%), followed by the Eastern (7.6%), Southern (6.9%), and Northern (5.7%) regions, and lowest in the Central region (2.8%).

Fig. 2 presents a choropleth map of WaSt prevalence across all governorates of Yemen. The map revealed substantial geographical variation, indicating high regional clustering of childhood undernutrition, with prevalence concentrated in high-burden areas — particularly in the northern and western parts of the country — rather than distributed evenly nationwide. Consistent with the regional estimates, the Western Coastal region showed the highest WaSt prevalence: the governorates of Al Hodeidah, Hajjah, Raymah, and Taiz, which constitute the western coastal belt, together with Lahj, were particularly affected, marking important hotspots for child undernutrition in the coastal corridor. The burden was comparatively moderate in the Northern region, although variation within the region was evident: Sa’dah and Al Jawf had comparatively high prevalence relative to other northern governorates such as Amran and Al Mahwit, indicating some clustering of risk in the northern belt. The Eastern region displayed a moderate WaSt burden, with governorates such as Hadramawt and Al Mahrah contributing to the regional load; although undernutrition persisted in these areas, it remained lower than in the western coastal zone. A similarly moderate prevalence pattern was observed in the Southern region, where nutritional vulnerability varied, with governorates such as Aden, Abyan, and Shabwah exhibiting higher levels of WaSt than others. The Central region had the lowest WaSt prevalence, largely driven by governorates that consistently fell into the lowest risk categories, including Sana’a City, Sana’a, Dhamar, and Al Bayda. Considered together, the map and the regional estimates indicate a distinct center-to-west gradient of childhood nutritional vulnerability in Yemen.

Fig. 3 displays a stacked column chart of WaSt prevalence and severe food insecurity by governorate, highlighting substantial geographical differences between nutritional outcomes and food access. The overall pattern shows that several governorates — including Al Jawf, Hajjah, Al Mahrah, and Sa’dah — had both high WaSt prevalence and high levels of severe food insecurity, indicating a high spatial overlap between acute malnutrition and food insecurity in these areas of high nutritional vulnerability. By contrast, urban and relatively stable governorates such as Sana’a City, Socotra, and Shabwah had comparatively lower WaSt prevalence and severe food insecurity, suggesting improved access to food systems and health services. Intermediate patterns, such as moderate food insecurity combined with moderate WaSt prevalence in governorates like Abyan, Lahj, Taiz, and Al Hodeidah, further illustrate the dose–response relationship between food insecurity and child undernutrition. Overall, the figure shows a strong positive spatial correlation between WaSt prevalence and severe food insecurity, indicating that concurrent wasting and stunting is more prevalent in governorates with higher levels of food insecurity.

**Fig. 3:**
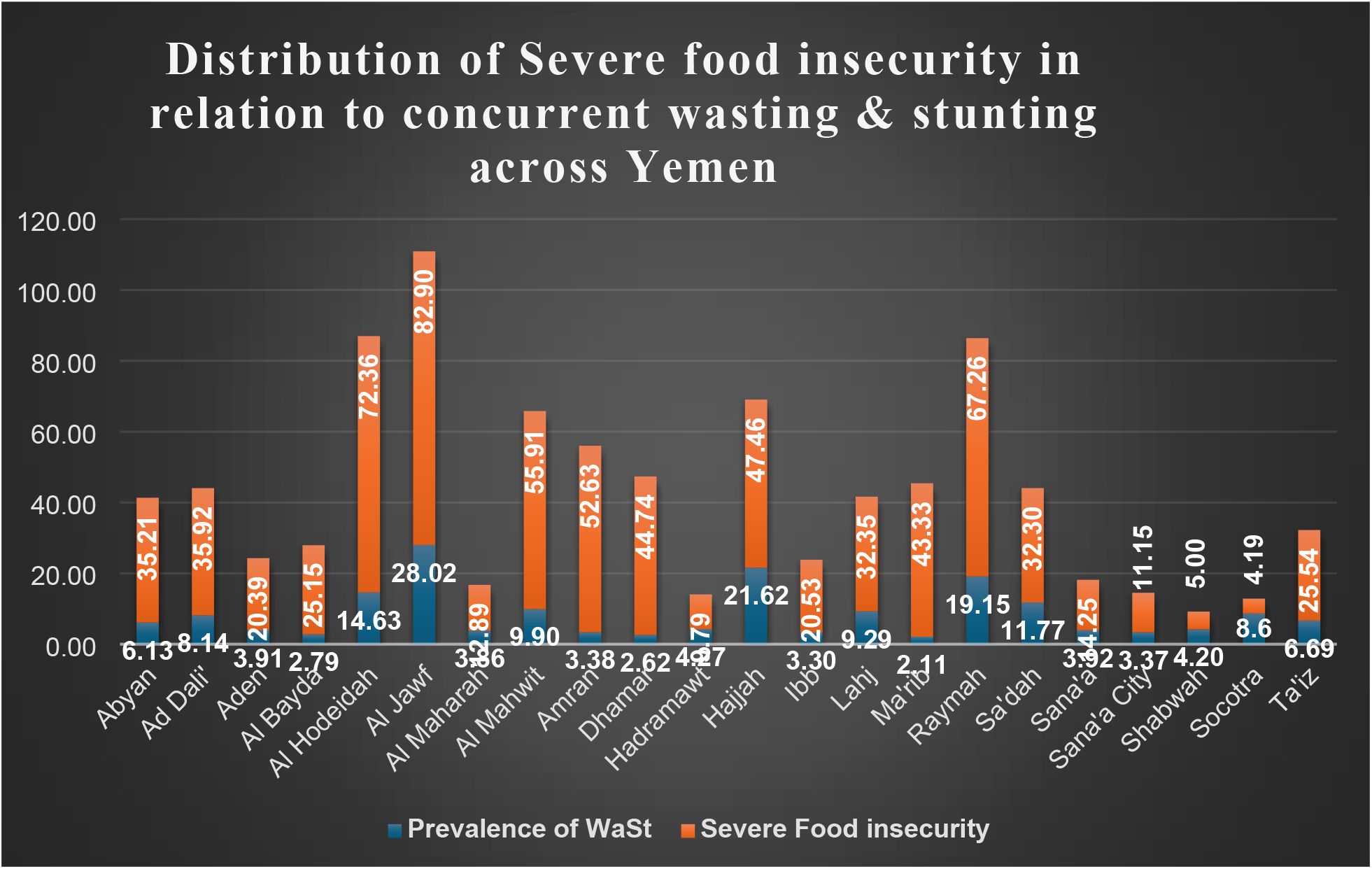
Stacked column chart showing Prevalence of severe food insecurity in relation to WaSt across all governorates in Yemen

### Multivariable logistic regression analysis of factors associated with WaSt

Table 2 presents the multivariable logistic regression analysis of factors associated with WaSt. In unadjusted analyses, child age, sex, area of residence, governorate region, household wealth index, food insecurity level, maternal education, maternal disability, recent fever, childhood diarrhoea, and WASH-related factors — including safe drinking water, sanitary toilet condition, and the presence of a designated handwashing area — were each significantly associated with WaSt (p < 0.05). After adjustment for potential confounders, child age, sex, household wealth index, food insecurity level, governorate region, maternal education, childhood diarrhoea, sanitary toilet condition, and the presence of a designated handwashing facility remained statistically significant predictors.

**Table 2:**
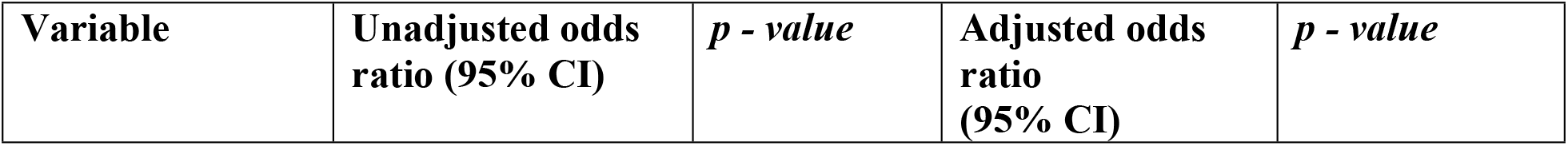
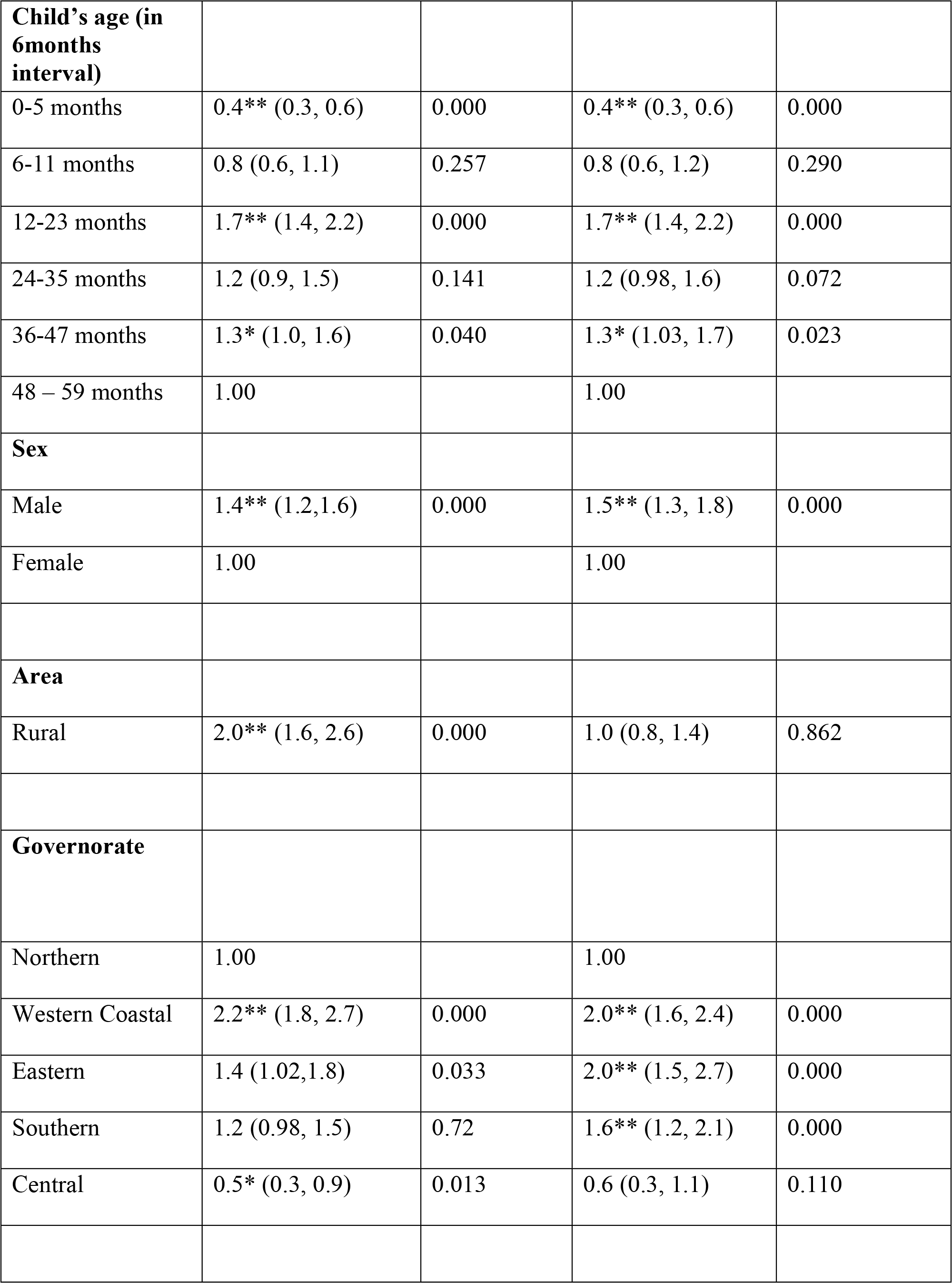
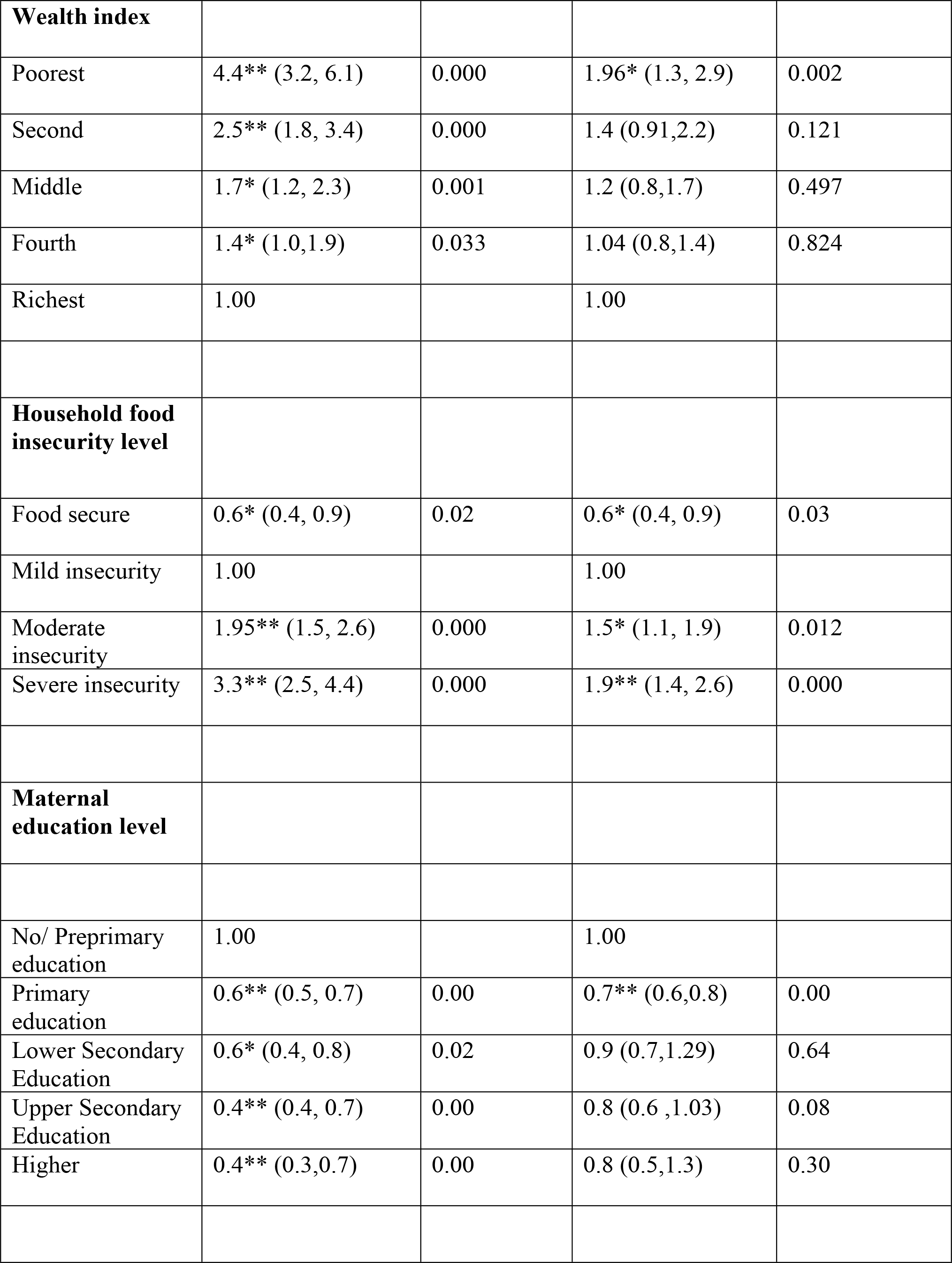
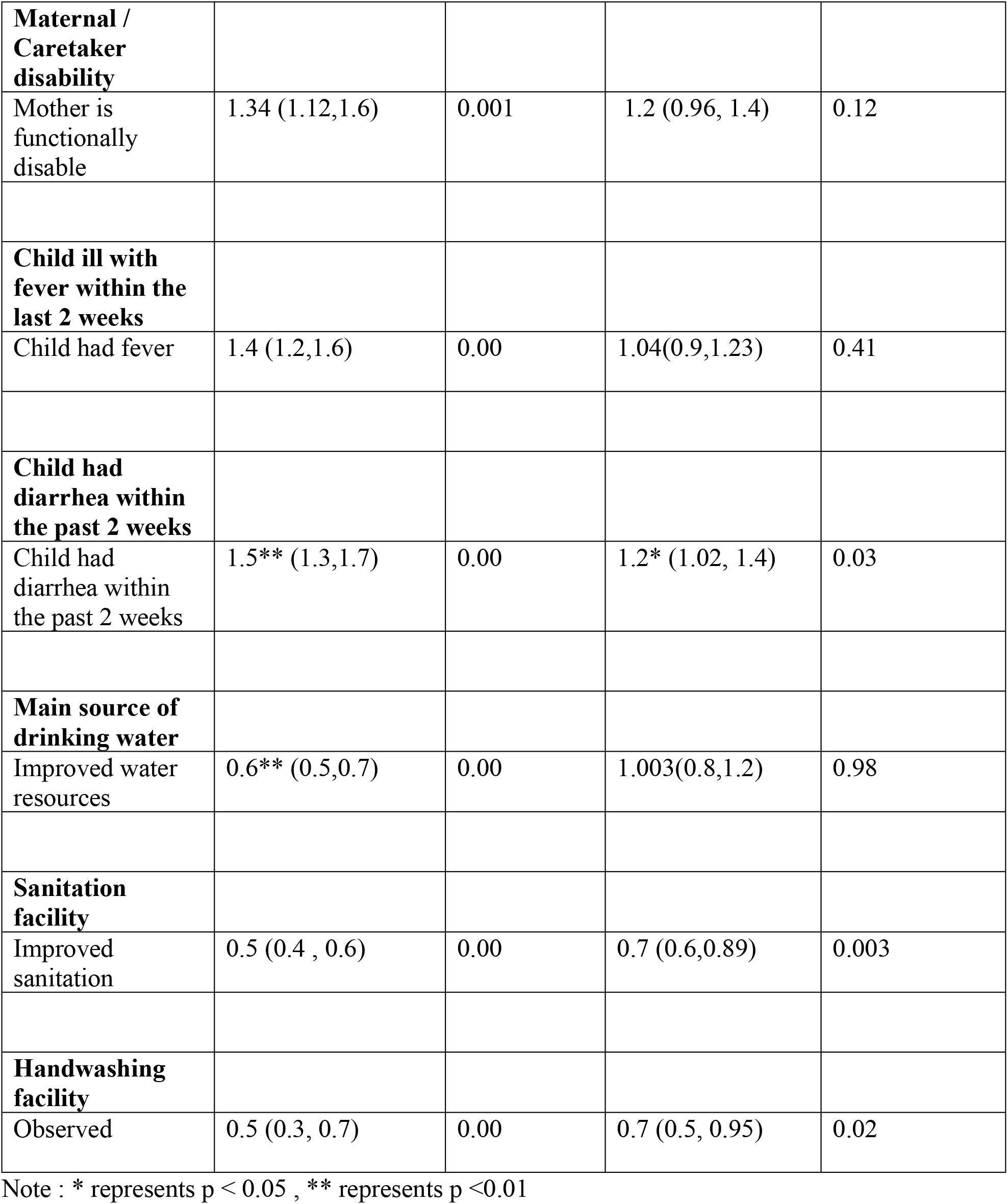
Multivariable logistic regression analysis of factors associated with concurrent wasting and stunting (WaSt) among under-five children.

Child age remained significantly associated with WaSt. Children were categorized into six age groups, with those aged 48–59 months serving as the reference group. Children aged 12–23 months had the highest odds of WaSt: concurrent wasting and stunting was 70% more common in this group than in the reference group (AOR = 1.70, 95% CI 1.4–2.2; p < 0.001). Children aged 36–47 months also had significantly higher odds (AOR = 1.3, 95% CI 1.03–1.7; p < 0.05), representing a 30% increase relative to the reference group. By contrast, infants aged 0–5 months had significantly lower odds of WaSt (AOR = 0.4, 95% CI 0.3–0.6; p < 0.001), a 60% reduction. No statistically significant associations were observed for children aged 6–11 or 24–35 months.

Child sex was significantly correlated with WaSt in the adjusted model: male children were 50% more likely than female children to experience WaSt (AOR = 1.52, 95% CI 1.3–1.8; p < 0.001). WaSt also varied significantly across Yemen’s five governorate regions, with the Northern region serving as the reference category. Children in the Western Coastal (AOR = 2.0, 95% CI 1.6–2.4; p < 0.001) and Eastern (AOR = 2.0, 95% CI 1.5–2.7; p < 0.001) regions had roughly twice the odds of WaSt of children in the North, and children in the Southern region had 60% higher odds (AOR = 1.6, 95% CI 1.20–2.10; p < 0.001). After adjustment, the association for children residing in the Central region was not statistically significant (p = 0.110).

The household wealth index remained a significant determinant of WaSt. Compared with children in the richest quintile, children in the poorest quintile were nearly twice as likely to be concurrently stunted and wasted (AOR = 1.96, 95% CI 1.3–2.9; p < 0.01), whereas no statistically significant associations were observed for the second, middle, or fourth wealth quintiles after adjustment.

Household food insecurity was substantially associated with WaSt, with mildly food-insecure households serving as the reference group. Children in moderately food-insecure households had 51% higher odds of WaSt (AOR = 1.51, 95% CI 1.13–1.94; p < 0.05), and children in severely food-insecure households were nearly twice as likely to have WaSt (AOR = 1.94, 95% CI 1.4– 2.6; p < 0.001). Conversely, children in food-secure households were 42% less likely to have WaSt than children in mildly food-insecure households (AOR = 0.58, 95% CI 0.35–0.95; p < 0.05).

With regard to maternal education, only primary education was significantly associated with WaSt: children whose mothers had completed primary school had 32% lower odds of WaSt (AOR = 0.68, 95% CI 0.56–0.84; p < 0.001) than children whose mothers had no or pre-primary education. Associations with lower secondary, upper secondary, and higher maternal education were not statistically significant after adjustment.

Children who had diarrhoea within the previous two weeks were 22% more likely to be stunted and wasted than those without diarrhoea (AOR = 1.22, 95% CI 1.02–1.44; p < 0.05). WaSt was 30% less common among children from households with improved sanitation than among those without (AOR = 0.70, 95% CI 0.56–0.89; p < 0.01), and the presence of a designated handwashing facility was associated with a 30% reduction in the odds of WaSt (AOR = 0.70, 95% CI 0.50–0.95; p < 0.05). However, use of an improved drinking water source was not independently associated with WaSt after adjustment. Similarly, maternal functional disability, urban or rural residence, and recent fever within the previous two weeks were not significantly associated with concurrent wasting and stunting in the adjusted model.

## Discussion

This study provides nationally representative evidence on the burden and determinants of concurrent wasting and stunting (WaSt) among under-five children in Yemen using MICS 2022– 2023 data. Among 17,734 children included in the final analysis, the overall prevalence of WaSt was 8.4%. This meant that approximately one in every 12 under-five children in Yemen was affected by both acute and chronic malnutrition. This finding indicates a substantial double burden of malnutrition in a fragile humanitarian context and highlights WaSt as an important public health concern requiring integrated both prevention and treatment strategies.

The present national analysis shows that WaSt is widespread across Yemen and shaped by various socioeconomic, geographic, food security, morbidity, and environmental determinants. The national prevalence observed in this study was slightly lower than the 10.7% prevalence reported among highly marginalized children in Sana’a city ^16^. This contrast suggests that WaSt is particularly concentrated in extremely deprived groups rather than being limited to marginalized settlements alone. While the Sana’a-based study provided a focused picture of WaSt under conditions of severe marginalization, this study extends the evidence by showing how structural inequalities operate across the wider population ^16^.

The burden of WaSt differed markedly by child sex and age. Male children had a higher WaSt prevalence than female children in bivariate analysis, and this association remained significant after adjustment, with boys having 50% higher odds of WaSt than girls. This finding is consistent with previous evidence from Yemen and other low- and middle-income settings showing male children are more at risk of early childhood malnutrition ^11,13,16,24–25^. The biological explanation for this phenomenon may include higher metabolic demands, greater vulnerability to infection, and sex-related differences in immune response ^26^. The persistence of this association even after adjustment with potential confounders suggests that male children may require closer nutritional monitoring, especially in high-risk households. This is consistent with recommendations to identify and reach children with concurrent wasting and stunting through existing nutrition programs ^13^.

Age was one of the strongest determinants for concurrent wasting and stunting found in the analysis. The highest WaSt prevalence was observed among children aged 12–23 months, and this group also had the highest adjusted odds of WaSt as well. Similar scenario was also seen in the study conducted among the highly marginalized children in Sana’a city ^16^. Compared with children aged 48–59 months, children aged 12–23 months had 70% higher odds of WaSt. This age pattern likely reflects increased vulnerability during the complementary feeding period, when children are exposed to inadequate dietary diversity, declining breastfeeding protection, contaminated foods, and repeated infections ^27^. Infants aged 0–5 months had significantly lower odds of WaSt, which may reflect the protective role of breastfeeding and lower exposure to contaminated complementary foods ^28^. Overall, these findings reinforce the importance of the first 1,000 days as the most critical window for preventing concurrent acute and chronic undernutrition.

This study showed that geographic disparities in concurrent wasting and stunting were substantial. WaSt prevalence varied significantly across Yemen’s regions, with the highest burden in the Western Coastal region, followed by the Eastern and Southern regions, while the Central region had the lowest prevalence. Previous studies have also shown that malnutrition in Yemen is strongly patterned by region, indicating that child undernutrition is not randomly distributed but concentrated in poorer and more vulnerable areas ^29^. In adjusted analysis, children living in the Western Coastal and Eastern regions had approximately two-fold higher odds of WaSt compared with those in the Northern region. And children in the Southern region also had 1.6 times higher odds which was statistically significant. The governorate-level choropleth map (shown in Fig 2) further demonstrated spatial clustering of WaSt, with high-burden areas including Al Hodeidah, Hajjah, Raymah, Taiz, Lahj, Sa’dah, Al Jawf, Hadramawt, and Al Mahrah. Similar findings were seen in the UNICEF Yemen 2025 nutrition report – “Nourishing the Future” , which highlighted that many women and children in specific areas of Yemen lack access to health care and preventive nutrition services, with large gaps in infant and young child feeding counselling and breastfeeding promotion ^30^. These patterns suggest that WaSt in Yemen is not randomly distributed, but concentrated in specific areas affected by food insecurity, conflict-related disruption, displacement, poor service access, and environmental vulnerability. Although WaSt prevalence was significantly higher in rural than urban areas in bivariate analysis, residence was no longer significant in the adjusted model. This was an interesting find in contrast to previous studies as they attributed this urban – rural difference of malnutrition to income inequality and structural regional disparities ^29^. But this study suggests that the apparent urban-rural difference may be explained by other factors, particularly region, household wealth, food insecurity, sanitation, and hygiene conditions. In Yemen’s context, urban residence does not necessarily indicate better living conditions, as many urban and peri-urban populations may face overcrowding, displacement, poor WASH access, and unstable food markets ^31^.

Persistent socioeconomic inequality among the households across all Yemen and its impact on WaSt was one of the clearest findings of this study. Children from the poorest households had almost two-fold higher odds of WaSt compared with those in the richest households. This shows that extreme poverty remains a major structural driver of concurrent wasting and stunting. This is a common finding also seen in the Sana’a city study ^16^. Interestingly in our analysis, intermediate wealth categories were not significant after adjustment, suggesting that the greatest excess risk is concentrated among the poorest households rather than distributed evenly across the socioeconomic gradient.

This study found that almost 36.2% of the households in the whole Yemen experience Severe Food insecurity followed by 41.4% of them experiencing moderate levels of Food insecurity. This finding puts Yemen at a much higher level of overall household food insecurity compared to previous studies, which reported that 22% of the households are severely food insecure ^32^. However, that particular study was based on only Hudaydah governorate , whereas this study worked on data from all the 22 governorates of Yemen. National IPC analyses indicate that approximately 50% of Yemen’s population has been classified in IPC Phase 3 or above (Crisis or worse) in recent years, with about one-third in Crisis (Phase 3) and an additional large proportion in Emergency (Phase 4) ^33^ . Our estimate aligns with these IPC findings, suggesting that a majority of households face at least moderate level of food insecurity.

During the bivariate and multivariate analysis, Food insecurity showed a strong and graded association with WaSt. WaSt prevalence increased progressively from food-secure to severely food-insecure households. Compared to households experiencing mild food insecurity level, children in the severely food-insecure households had almost twice the odds of WaSt. Conversely, food security at the household level had a significant protective effect against concurrent wasting and stunting. Children who hailed from Food secure houses had 40% less likelihood of experiencing WaSt. A previously done systematic review and meta-analysis of household food insecurity and stunting reported that moderate and severe food insecurity tended to be associated with higher risks of stunting compared with food-secure or mildly insecure households, even though pooled estimates were attenuated by heterogeneity ^34^. A more comprehensive study on household food insecurity and it’s link with child undernutrition found that children living in food-insecure households had significantly greater odds of experiencing wasting, stunting, and underweight, with the risk increasing as the severity of food insecurity worsened ^35^. This dose-response pattern strongly supports the role of household food insecurity as a proximal pathway linking Yemen’s humanitarian crisis, economic instability, and disrupted food systems to child growth failure. A clear picture of how food insecurity is raising WaSt across the 22 governorates of Yemen was painted in the stacked bar chart (shown in the Fig 3). The spatial overlap between severe food insecurity and WaSt across governorates further strengthens this interpretation.

This study showed that maternal education had a protective but a partially attenuated association. Children of mothers with no or pre-primary education had the highest WaSt prevalence, while those whose mothers had higher education had the lowest prevalence in bivariate analysis. Similar findings had been seen on previously conducted systematic reviews as well ^36^. However, after adjustment, only primary maternal education remained significantly protective against WaSt. Compared with children whose mother had no or pre-primary education, the children of mothers who had at least primary education had 32% less odds of having concurrent wasting and stunting. This finding is supported by previously conducted similar studies based on DHS data ^37^. A study based on the urban cohort of South Africa indicated that maternal education exerted an indirect influence on stunting, primarily through socio-economic status and the antenatal environment, as reflected by the birth weight Z-score ^38^. This explains the loss of significance for higher education categories in this crisis-affected settings of Yemen. Because the effect of education is constrained by household poverty, food insecurity, and environmental deprivation. Even so, robust evidences have declared that lower levels of maternal and paternal education remained significant risk factors for child mortality after adjusting for other indicators of family socioeconomic status ^39^. These results provide strong evidence that promoting universal access to quality education may contribute to achieving Sustainable Development Goal (SDG) target 3.2, which aims to reduce neonatal and under-five mortality.

In the present study, 38.3% of children under five years of age experienced diarrhea during the two weeks preceding the survey. This prevalence was comparable to the 37.3% reported by Satty et al. (2025), who analyzed the same Yemen MICS dataset ^22^. The slight difference in prevalence may be attributable to variations in the study populations, as Satty et al. included only children aged 6–59 months, whereas the present study examined children aged 0–59 months (see fig 1: participant flow diagram) . In this study, the prevalence of WaSt among the diarrhea affected children is significantly much higher compared to the group that has not experienced diarrhea in the last 2 weeks. Moreover in the multivariate analysis, Recent (within <= 2 weeks) childhood diarrhea was an important morbidity-related determinant. Children who had diarrhea within the previous two weeks had 22% higher odds of WaSt compared with children without diarrhea. This finding is very consistent with the infection-malnutrition cycle, where diarrheal illness reduces nutrient absorption, worsens appetite, and contributes to weight loss, while malnourished children become more susceptible to infection described in the study by Guerrant et al. in 2003^40^. A study based on the Diarrheal Disease Surveillance system (DDSS) in icddrb, Bangladesh found that children with concourrent severe wasting and severe stunting more often present with dehydrating diarrhoea than those with either condition alone, similar to this analysis ^41^. In the Sana’a marginalized community study, diarrhea was also a common determinant of WaSt ^16^. The consistent association between diarrhea and WaSt in both this national analysis and the Sana’a marginalized-community study reinforce the importance of preventing enteric infections as part of WaSt reduction strategies.

As diarrheal diseases are closely related to WASH practices, the condition of drinking water, condition of sanitary toilet and handwashing area was considered in this study. Although majority of the households had access to safe drinking water (79.2%), improved sanitation (82.1%) and designated handwashing area (89.8%), these WASH related determinants played a significant role in the prevention of WaSt, as seen in the multivariable logistic regression analysis. The children whose households had improved sanitation, the odds of WaSt were 30% less among them. Also having a designated handwashing area was independently protective. There was 30% less odds of WaSt among the children whose households had a designated handwashing area. Previous study by Cameron et al. based on the data from Indonesian Family Life Survey(IFLS) reported that improved household sanitation was associated with around a 5 percentage - point reduction (equivalent to 30% reduction) in childhood stunting and highlighted that benefits increase as sanitation coverage approaches universality ^42^. The Global Handwashing Partnership has stated that based on WHO data, about 50% of childhood undernutrition is attributable to repeated diarrhea and intestinal infections from poor WASH practices ^43^. The evidence further suggests that clean water and handwashing with soap can reduce stunting by up to 15% by preventing nutrient loss through diarrhea ^43^.

Some variables that were significant in bivariate analysis lost significance after adjustment. Recent fever, Maternal functional disability, urban-rural residence, agricultural land ownership, domestic animal ownership, and improved water sources were not independently associated with WaSt in the final model. This suggests that their crude associations may be explained by underlying socioeconomic, regional, food security, and WASH-related factors. These findings highlight the importance of multivariable analysis in distinguishing direct predictors from variables that act through indirect broader pathways.

Overall, this study shows that WaSt among under-five children in Yemen is driven by a multi-layered interaction of biological vulnerability, poverty, food insecurity, infection, geographic location, and environmental deprivation. The findings from the Sana’a marginalized-community study and the present national analysis are complementary. Both identify male sex, a household wealth quantile and diarrheal illness as consistent risk factors ^16^, while the national analysis additionally reveals the dominant role of various other household factors such as food insecurity, regional clustering, maternal education, sanitation, and handwashing access. This study also points out that the specific age group between 12-23 months is the most vulnerable period for concurrent wasting and stunting. These findings suggest that concurrent wasting and stunting in Yemen should be viewed not merely as a clinical or nutritional problem, but as a consequence of overlapping between structural inequalities and humanitarian deprivations.

## Recommendations

The public health implications based on the findings are clear. Concurrent wasting and stunting is a real burning threat to both the present and future of Yemen, ranking in almost on the top of its challenges. To solve this mess, smart and precise interventions are a timely need. These interventions should prioritize the poorest and most food-insecure households, and high-burden regions such as the Western Coastal, Eastern, and Southern areas. During community-based screening and awareness programs, particular attention should be given to male children and those who have entered the complementary feeding period, as they may be at greater risk of concurrent wasting and stunting. Previously , icddr,b reported that malnutrition damages healthy gut bacteria in children, reinforcing that repeated enteric infections and poor diets impair intestinal absorptive function and contribute to growth faltering ^44^. To address this challenge, scientists at icddr,b developed microbiota-directed complementary food (MDCF-2) and evaluated it in a clinical trial ^45^. Among Bangladeshi children with moderate acute malnutrition, MDCF-2 resulted in greater weight gain than conventional ready-to-use supplementary food (RUSF), while also promoting maturation of the gut microbiota and improving biomarkers of intestinal health and environmental enteric dysfunction ^46^.Given the high mortality risk and frequent environmental enteric dysfunction among children with WaSt, microbiota-directed complementary foods (MDCF-2) may complement standard nutritional management by improving nutrient absorption and gut health ^47^ . Therefore, nutritional programs can ensure supply of MDCF-2 to the severely affected communities of Yemen and reduce the double burden of wasting and stunting. However, programs should combine nutrition-specific interventions with food security support, diarrheal disease prevention, sanitation improvement, handwashing promotion, and strengthened primary healthcare services. In Yemen’s context, isolated treatment of wasting or stunting is unlikely to be sufficient. WaSt requires integrated, equity-focused, and geographically targeted strategies that address the shared pathways linking acute and chronic undernutrition.

## Limitations and strengths

This study has some mentionable limitations as well. First, the cross-sectional design of the 2022 Yemen Multiple Indicator Cluster Survey (MICS) precludes causal inference between the identified factors and concurrent wasting and stunting (WaSt). Secondly, the use of secondary data limited the analysis to variables only available in the dataset. In addition, the caregiver-reported information on childhood illnesses and household food insecurity may be subject to recall and reporting bias. Moreover, the aggregation of governorates into five regions may have masked subregional heterogeneity. Despite these limitations, the study utilized a large, nationally representative sample and survey-weighted analyses accounting for the complex MICS sampling design, enhancing the generalizability of the findings to under-five children in Yemen.

## Conclusion

In conclusion, concurrent wasting and stunting among under-five children in Yemen represents a serious public health problem rooted in poverty, food insecurity, infection, poor sanitation, and regional vulnerability. The national prevalence of 8.4% confirms that WaSt is not only restricted to marginalized settlements. Truth be told, it is intensified among the most deprived populations. Preventing WaSt in Yemen will require early screening at the community level, smart early-life interventions such as complementary food supply through comprehensive nutritional programs, improved household food security, stronger WASH systems, and targeted support for geographically vulnerable communities.

## Author contribution

AS – Conceptualization , methodology , data analysis , interpretation , manuscript writing ; RS – data interpretation , draft manuscript writing and review ; BK – draft manuscript writing and critical review ; RDG – contributed as the senior author in supervision, methodology , data analysis plan, critical review and conceptualization.

## Funding

This research did not receive any specific funding or support by any organization. Every part of the research was carried out by the authors personal effort and skills.

## Data Availability

Access to MICS datasets is available for free by submitting a request on the MICS website . Link of the survey dataset website : https://mics.unicef.org/surveys?display=card&keys=Yemen. The data was accessed on 11 January, 2026 by the authors.

## Declarations

### Ethics approval and consent to participate

Data was accessed after formally registering for research on the MICS website (https://mics.unicef.org/surveys?display=card&keys=Yemen). The publicly available Yemen MICS 2022-2023 data set was anonymized and de-identified to safeguard the confidentiality of respondents.

### Consent for publication

The authors confirm that they have all reviewed and approved the manuscript for publication.

### Competing interests

The authors declare no competing interests.

## Notes

### Competing Interest Statement

The authors have declared no competing interest.

### Author Declarations

This study was a secondary analysis of the Yemen Multiple Indicator Cluster Survey (MICS) 2022–2023 dataset. The dataset was obtained through the official MICS data-access process following registration for research purposes. The dataset available to the authors was anonymized and de-identified, and no personally identifiable information was accessed. As this study involved only secondary analysis of existing anonymized data and did not involve recruitment or direct interaction with human participants, no additional institutional ethical approval or informed consent was required for this secondary analysis.

## References

1) UNICEF. Malnutrition [Internet]. New York: UNICEF; [cited 2026 Jul 13]. Available from: https://data.unicef.org/topic/nutrition/malnutrition/

2) Verma P, Prasad JB. Stunting, wasting and underweight as indicators of under-nutrition in under five children from developing Countries: A systematic review. Diabetes & Metabolic Syndrome: Clinical Research & Reviews. 2021 Sep 1;15(5):102243.

3) World Health Organization. Child malnutrition: Wasting among children under 5 years of age [Internet]. Geneva: World Health Organization; [cited 2026 Jul 13]. Available from: https://www.who.int/data/gho/indicator-metadata-registry/imr-details/302

4) World Health Organization. Global nutrition targets 2030: stunting policy brief [Internet]. Geneva: World Health Organization; 2025 [cited 2026 Jul 13]. Available from: https://www.who.int/publications/i/item/B09383

5) De Sanctis V, Soliman A, Alaaraj N, Ahmed S, Alyafei F, Hamed N. Early and Long-term Consequences of Nutritional Stunting: From Childhood to Adulthood. Acta Biomed. 2021 Feb 16;92(1):e2021168. doi: 10.23750/abm.v92i1.11346. PMID: 33682846; PMCID: PMC7975963.

6) Mertens A, Benjamin-Chung J, Colford JM Jr, Hubbard AE, van der Laan MJ, Coyle J, Sofrygin O, Cai W, Jilek W, Rosete S, Nguyen A, Pokpongkiat NN, Djajadi S, Seth A, Jung E, Chung EO, Malenica I, Hejazi N, Li H, Hafen R, Subramoney V, Häggström J, Norman T, Christian P, Brown KH, Arnold BF; Ki Child Growth Consortium. Child wasting and concurrent stunting in low- and middle-income countries. Nature. 2023 Sep;621(7979):558–567. doi: 10.1038/s41586-023-06480-z. Epub 2023 Sep 13. Erratum in: Nature. 2023 Nov;623(7985):E1. doi: 10.1038/s41586-023-06695-0. PMID: 37704720; PMCID: PMC10511327.

7) Zaba, T., Conkle, J., Nyawo, M. et al. Concurrent wasting and stunting among children 6–59 months: an analysis using district-level survey data in Mozambique. BMC Nutr 8, 15 (2022). 10.1186/s40795-022-00508-9

8) Myatt M, Khara T, Schoenbuchner S, Pietzsch S, Dolan C, Lelijveld N, Briend A. Children who are both wasted and stunted are also underweight and have a high risk of death: a descriptive epidemiology of multiple anthropometric deficits using data from 51 countries. Arch Public Health. 2018 Jul 16;76:28. doi: 10.1186/s13690-018-0277-1. PMID: 30026945; PMCID: PMC6047117.

9) Saaka M, Galaa SZ. Relationships between Wasting and Stunting and Their Concurrent Occurrence in Ghanaian Preschool Children. J Nutr Metab. 2016;2016:4654920. doi: 10.1155/2016/4654920. Epub 2016 Jun 9. PMID: 27379184; PMCID: PMC4917721.

10) Prendergast, A. J., & Humphrey, J. H. (2014). The stunting syndrome in developing countries. Paediatrics and International Child Health, 34(4), 250–265. 10.1179/2046905514Y.0000000158

11) Odei Obeng-Amoako GA, Karamagi CAS, Nangendo J, et al. Factors associated with concurrent wasting and stunting among children 6–59 months in Karamoja, Uganda. Matern Child Nutr. 2021; 17:e13074. 10.1111/mcn.13074

12) World Health Organization. Joint child malnutrition estimates (UNICEF-WHO-WB) [Internet]. Geneva: World Health Organization; [cited 2026 Jul 20]. Available from: https://www.who.int/data/gho/data/themes/topics/joint-child-malnutrition-estimates-unicef-who-wb

13) Khara T, Mwangome M, Ngari M, Dolan C. Children concurrently wasted and stunted: A meta-analysis of prevalence data of children 6-59 months from 84 countries. Matern Child Nutr. 2018 Apr;14(2):e12516. doi: 10.1111/mcn.12516. Epub 2017 Sep 25. PMID: 28944990; PMCID: PMC5901398.

14) United Nations Children’s Fund (UNICEF). Yemen crisis [Internet]. New York: UNICEF; [cited 2026 Jul 20].

15) United Nations. Yemen: Children continue to bear the brunt of conflict as humanitarian needs soar [Internet]. New York: United Nations; 2025 Mar [cited 2026 Jul 20]. Available from: https://news.un.org/en/story/2025/03/1161471

16) Al-Taj MA, Al Serouri A, Al-Muradi AM, Al-Dharhani E, Al-Faeq NN, Al-Amodi FM, Abdulwahab MM, Nawfal AM, Alshemerry MH, Mujahed MA. Concurrent wasting and stunting among marginalised children in Sana’a city, Yemen: a cross-sectional study. J Nutr Sci. 2023 Aug 4;12:e91. doi: 10.1017/jns.2023.72. PMID: 37587974; PMCID: PMC10425760.

17) Central Statistical Organization (CSO) and United Nations Children’s Fund (UNICEF). Yemen multiple Indicator Cluster Survey 2022–2023, Survey findings Report. Yemen: Central Statistical Organization and United Nations Children’s Fund; 2023.

18) Khan S, Hancioglu A. Multiple Indicator Cluster Surveys: Delivering Robust Data on Children and Women across the Globe. Stud Fam Plann. 2019 Sep;50(3):279–286. doi: 10.1111/sifp.12103. Epub 2019 Sep 5. PMID: 31486080; PMCID: PMC6771654.

19) World Bank. What is food security? There are four dimensions [Internet]. Washington (DC): World Bank; [cited 2026 Jul 21]. Available from: https://www.worldbank.org/en/topic/agriculture/brief/food-security-update/what-is-food-security

20) Food and Agriculture Organization of the United Nations. Applying the Food Insecurity Experience Scale (FIES) [Internet]. Rome: FAO; [cited 2026 Jul 21]. Available from: https://www.fao.org/measuring-hunger/access-to-food/applying-the-fies/en

21) Mahmood T, Kumar R, Ali TM, Naeem N, Pongpanich S. Determinants of the food insecurity at household level in Pakistan: A multilevel model approach. Plos one. 2023 Oct 5;18(10):e0291343.

22) Satty A, Salih M, Abdalla FA, A Mahmoud AF, Gumma EAE, Saad Mohamed Khamis G, Adam AMA, Hassaballa AA, Hamed OMA, M S Mohammed Z. Statistical Analysis of Factors Associated with Diarrhea in Yemeni Children under Five: Insights from the 2022-2023 Multiple Indicator Cluster Survey. J Epidemiol Glob Health. 2024 Sep;14(3):1043-1051. doi: 10.1007/s44197-024-00253-1. Epub 2024 Jun 10. Erratum in: J Epidemiol Glob Health. 2024 Sep;14(3):1368. doi: 10.1007/s44197-024-00268-8. PMID: 38856818; PMCID: PMC11442882.

23) Hosmer D, Lemeshow S. Applied Logistic Regression. 2nd ed. Wiley; 2000.

24) Schoenbuchner SM, Dolan C, Mwangome M, Hall A, Richard SA, Wells JC, Khara T, Sonko B, Prentice AM, Moore SE. The relationship between wasting and stunting: a retrospective cohort analysis of longitudinal data in Gambian children from 1976 to 2016. Am J Clin Nutr. 2019 Aug 1;110(2):498–507. doi: 10.1093/ajcn/nqy326. PMID: 30753251; PMCID: PMC6669055.

25) Thurstans S, Opondo C, Seal A, Wells J, Khara T, Dolan C, Briend A, Myatt M, Garenne M, Sear R, Kerac M. Boys are more likely to be undernourished than girls: a systematic review and meta-analysis of sex differences in undernutrition. BMJ Glob Health. 2020 Dec;5(12):e004030. doi: 10.1136/bmjgh-2020-004030. PMID: 33328202; PMCID: PMC7745319.

26) Thompson AL. Greater male vulnerability to stunting? Evaluating sex differences in growth, pathways and biocultural mechanisms. Ann Hum Biol. 2021 Sep;48(6):466–473. doi: 10.1080/03014460.2021.1998622. PMID: 35105202; PMCID: PMC9205267.

27) Aboagye RG, Seidu AA, Ahinkorah BO, Arthur-Holmes F, Cadri A, Dadzie LK, Hagan JE Jr, Eyawo O, Yaya S. Dietary Diversity and Undernutrition in Children Aged 6-23 Months in Sub-Saharan Africa. Nutrients. 2021 Sep 28;13(10):3431. doi: 10.3390/nu13103431. PMID: 34684435; PMCID: PMC8537414.

28) World Health Organization. Breastfeeding: exclusive breastfeeding under 6 months (%). Geneva: World Health Organization; c2024 [cited 2026 Jul 6]. Available from: https://www.who.int/data/gho/indicator-metadata-registry/imr-details/130

29) Sharaf M, Rashad AS. Regional inequalities in child malnutrition in Egypt, Jordan and Yemen: a Blinder–Oaxaca decomposition analysis. Health Econ Rev. 2016;6(1):23.

30) United Nations Children’s Fund (UNICEF) Yemen. Nourishing the future: Investing in nutrition for child development and nation-building in Yemen [Internet]. Sana’a (Yemen): UNICEF Yemen; 2025 Mar [cited 2026 Jul 6]. Available from: https://www.unicef.org/yemen/reports/nourishing-future-0

31) Action Against Hunger. In Yemen, the population is struggling to meet its basic needs [Internet]. Paris: Action Against Hunger; 2024 Mar 26 [cited 2026 Jul 6].

32) Perez Duque M, Alburhomy A, Ahmed A, Saleh Y, Wolz A, B. Abecasis A, J. Saad Duque N, Salje H, Gil-Cuesta J, Robays J, Veicht R. Acute malnutrition and food insecurity in Yemen, 2021: Evidence from a two-stage cluster randomised survey in a protracted crisis. PLOS Global Public Health. 2025 Jul 11;5(7):e0004331.

33) Food and Agriculture Organization of the United Nations. Conflict and economic crisis drive food insecurity and malnutrition in Yemen [Internet]. Rome: FAO; 2023 May 25 . Available from: https://www.fao.org/newsroom/detail/conflict-and-economic-crisis-drive-food-insecurity-and-malnutrition-in-yemen/

34) Patriota ÉS, Abrantes LC, Figueiredo AC, Pizato N, Buccini G, Gonçalves VS. Association between household food insecurity and stunting in children aged 0− 59 months: Systematic review and meta-analysis of cohort studies. Maternal & child nutrition. 2024 Apr;20(2):e13609.

35) Sutrisno R, Fatmawati ID, Laksono AD, Rahmawati W, Wirawan NN, Handayani D. The role of household food insecurity in malnutrition among Indonesian children under 5 years of age: a systematic review and meta-analysis (2015-2025). Public Health Nutr. 2026 Mar 26;29(1):e86. doi: 10.1017/S1368980026102365. PMID: 41883328; PMCID: PMC13112310.

36) Rezaeizadeh G, Mansournia MA, Keshtkar A, Farahani Z, Zarepour F, Sharafkhah M, Kelishadi R, Poustchi H. Maternal education and its influence on child growth and nutritional status during the first two years of life: a systematic review and meta-analysis. EClinicalMedicine. 2024 Apr 4;71:102574. doi: 10.1016/j.eclinm.2024.102574. PMID: 38596614; PMCID: PMC11001623.

37) Lawal SA, Okunlola DA, Adegboye OA, Adedeji IA. Mother’s education and nutritional status as correlates of child stunting, wasting, underweight, and overweight in Nigeria: Evidence from 2018 Demographic and Health Survey. Nutrition and Health. 2024 Oct;30(4):821–30.

38) Casale D, Espi G, Norris SA. Estimating the pathways through which maternal education affects stunting: evidence from an urban cohort in South Africa. Public health nutrition. 2018 Jul;21(10):1810–8.

39) Balaj M, York HW, Sripada K, Besnier E, Vonen HD, Aravkin A, Friedman J, Griswold M, Jensen MR, Mohammad T, Mullany EC, Solhaug S, Sorensen R, Stonkute D, Tallaksen A, Whisnant J, Zheng P, Gakidou E, Eikemo TA. Parental education and inequalities in child mortality: a global systematic review and meta-analysis. Lancet. 2021 Aug 14;398(10300):608–620. doi: 10.1016/S0140-6736(21)00534-1. Epub 2021 Jun 10. PMID: 34119000; PMCID: PMC8363948.

40) Guerrant RL, Oriá RB, Moore SR, Oriá MO, Lima AA. Malnutrition as an enteric infectious disease with long-term effects on child development. Nutr Rev. 2008 Sep;66(9):487–505. doi: 10.1111/j.1753-4887.2008.00082.x. PMID: 18752473; PMCID: PMC2562291.

41) Nuzhat S, Shahunja KM, Shahid ASMSB, Khan SH, Islam SB, Islam MR, Ahmed T, Chisti MJ, Hossain MI, Faruque ASG. Diarrhoeal children with concurrent severe wasting and stunting compared to severe wasting or severe stunting. Trop Med Int Health. 2020 Aug;25(8):928–935. doi: 10.1111/tmi.13446. Epub 2020 Jun 15. PMID: 32446268.

42) Cameron L, Chase C, Haque S, Joseph G, Pinto R, Wang Q. Childhood stunting and cognitive effects of water and sanitation in Indonesia. Economics & Human Biology. 2021 Jan 1;40:100944.

43) Global Handwashing Partnership. Nutrition [Internet]. Washington (DC): Global Handwashing Partnership; [cited 2026 Jul 13]. Available from: https://globalhandwashing.org/about-handwashing/why-handwashing/nutrition/

44) Subramanian S, Huq S, Yatsunenko T, Haque R, Mahfuz M, Alam MA, Benezra A, DeStefano J, Meier MF, Muegge BD, Barratt MJ, VanArendonk LG, Zhang Q, Province MA, Petri WA Jr, Ahmed T, Gordon JI. Persistent gut microbiota immaturity in malnourished Bangladeshi children. Nature. 2014 Jun 19;510(7505):417–21. doi: 10.1038/nature13421. Epub 2014 Jun 4. PMID: 24896187; PMCID: PMC4189846.

45) icddr,b. MDCF-2: Mobilising the gut microbiome to tackle undernutrition [Internet]. Dhaka: International Centre for Diarrhoeal Disease Research, Bangladesh; 2025 Oct 12 [cited 2026 Jul 22]. Available from: https://www.icddrb.org/features/mdcf-2-mobilising-the-gut-microbiome-to-tackle-undernutrition-12-10-2025

46) Mostafa I, Lamiya UH, Rasul MG, Naila NN, Fahim SM, Hasan SMT, Barratt MJ, Gordon JI, Ahmed T. Development and Acceptability of Shelf-Stable Microbiota Directed Complementary Food Formulations. Food Nutr Bull. 2024 Jun;45(2-3):67–73. doi: 10.1177/03795721241250104. Epub 2024 Jul 30. PMID: 39077991.

47) Mostafa I, Sthity RA, Lamiya UH, Tariqujjaman M, Mahfuz M, Hasan SMT, Ahmed T. Effect of Gut Microbiota-Directed Complementary Food Supplementation on Fecal and Plasma Biomarkers of Gut Health and Environmental Enteric Dysfunction in Slum-Dwelling Children with Moderate Acute Malnutrition. Children (Basel). 2024 Jan 6;11(1):69. doi: 10.3390/children11010069. PMID: 38255381; PMCID: PMC10814735

